# Enabling reproducible type 1 diabetes polygenic risk scoring for clinical and translational applications

**DOI:** 10.1101/2025.07.15.25331523

**Authors:** Maizy S. Brasher, Matthew J. Fisher, Carolina Sanchez Wild, Jonathan A. Shortt, Krista Miller, Randi K. Johnson, Nicholas M. Rafaels, Elizabeth L. Kudron, Ian M. Brooks, Kristy R. Crooks, Sean M. Oser, Tamara K. Oser, Joanne B. Cole, Colorado Center for Personalized Medicine, Laura K. Wiley, Sridharan Raghavan, Neda Rasouli, Meng Lin, Christopher R. Gignoux

**Author notes:** These authors contributed equally (Meng Lin, Christopher R. Gignoux).

## Abstract

**Objective:** Type 1 diabetes polygenic risk scores (PRS) offer a promising tool for identifying diabetes subtypes in adults with new-onset disease. We aimed to develop a pipeline for the clinical translation of type 1 diabetes PRS to support clinical decision-making within a large health system and to provide publicly available code for applying these methods to future PRS models.

**Research Design and Methods:** We adapted two established type 1 diabetes PRS models: a 67-SNP (GRS2) and a 7-SNP (AA7) score for a clinical genotyping platform and applied them to 73,346 participants in the biobank at the Colorado Center for Personalized Medicine (CCPM). We evaluated the scores’ performance differentiating between type 1 and type 2 diabetes in adults using a clinician-curated diabetes phenotyping algorithm and examined associations with diabetes-related clinical data extracted from patients’ health records. The impact of technical genotyping missingness on score accuracy and ancestry calibration were assessed independently.

**Results:** Both scores effectively distinguished type 1 from type 2 diabetes across genetically defined ancestry groups (all AUC > 0.80) and demonstrated consistent performance in the UK Biobank (all AUC > 0.75). Individuals in the top quintile of each PRS were enriched for diabetic ketoacidosis (DKA) cases, accounting for nearly half of all DKA cases in the cohort. Additionally, the top quintile showed nearly threefold increased odds of GAD autoantibody positivity (OR = 2.94 [95% CI 2.08-4.17]).

**Conclusions:** Our evaluations demonstrated the potential utility of PRS for diabetes subtyping in a clinical setting. We present a framework of critical steps toward a standardized system for future translation of diabetes PRS to equitable clinical use, along with software to make it possible for others.

## Introduction

Polygenic scores are becoming increasingly common in complex trait genetics as they allow researchers to quantify genetic predispositions by aggregating the effects of thousands of genetic variants. Advances in genome-wide association studies (GWAS) and computational methods have significantly improved the accuracy and applicability of these scores. As a result, polygenic scores are now increasingly being integrated into research on disease risk prediction and touted for their potential for much greater sensitivity in personalized medicine than traditional clinical genetic approaches^1^.

Type 1 diabetes remains a key example of polygenic scoring due to the high prevalence (4-6% among diabetes cases in the United States^2–4^), a strong genetic component (heritability around 50%^5^), and strong effect sizes present in the genome^6,7^. It can be used for risk stratification, subtyping, and recommending changes in behavior and/or medication use to avoid potentially life-threatening complications. And with the advent of novel prevention therapies such as teplizumab, there is an opportunity to prevent or delay the onset of clinical manifestations if identified in a preclinical state. This is particularly relevant in the adult setting: Adult-onset autoimmune diabetes (commonly called Latent Autoimmune Diabetes in Adults, LADA) shares a similar mechanism to type 1 diabetes, yet it is often misdiagnosed or misannotated as type 2 diabetes due to later age of onset and co-occurrence with clinical risk factors like obesity. While autoantibody testing is an important confirmatory approach for type 1 diagnosis, it may not be ordered at the early stage; changes in antibody presentation at older ages can complicate subtype diagnosis^8–10^. As a potential solution, an affordable clinical PRS test that can aid in differentiating between type 1 diabetes and type 2 diabetes could result in low-risk, high-impact clinical interventions and changes to the standard of care, such as higher frequency of autoantibody testing or changes to insulin management. There has been increasing evidence of the importance of autoimmune diabetes beyond the juvenile-onset setting^6,11^.

While PRS shows a lot of promise, there remain significant barriers to clinical and translational applications. These are partially mitigated in a type 1 diabetes setting. A primary technical hinderance results from the extensive number of variants used in many scores, often ranging from tens of thousands to millions from development using whole-genome sequencing or imputed genetic data. The number far exceeds the feasibility for clinical implementation due to the prohibitive time and effort required to comply with analytic validity and regulatory requirements. However, this is less of an issue in the T1D setting. Multiple type 1 diabetes scores have shown high prediction accuracy only using tens to hundreds of variants^7,12^.

Some other barriers persist with type 1 diabetes PRSs. First, people may not have access to a platform with perfect coverage of all variants matching a published score. While genome-wide imputation may help, it may not be a clinically valid technique due to imperfect accuracy and regulatory concerns. Second, genetic ancestry is widely known to be a major confounder in polygenic scoring. While multiple investigations have shown the utility of PRS in a multi-ancestry setting (e.g. https://doi.org/10.2337/dc25-0142) these typically focus on retrospective, population-stratified analyses. This has limited translation into the clinic where a score should work for anyone who walks into a clinic, particularly for less well-represented groups or individuals with multiple ancestries^13^. Third, there is need for a systematic approach to the evaluation and implementation of PRS in translational and clinical settings^14^, demonstrating PRS’s additional value alongside existing clinical variables. Importantly, the problems of imperfect coverage and ancestry confounding are likely to be issues across current and future polygenic scores.

To address these concerns and bring type 1 diabetes PRS closer to clinical and translational applications, we present a path to clinical translation of PRS for diabetes classification, with possible intervention and opportunities for clinical decision support and additional diagnostic/treatment recommendations in a large health system. Importantly, we develop a framework for tag SNP selection and ancestry recalibration and provide software for both, enabling researchers with heterogeneous data to perform robust, high-quality T1D polygenic scoring using existing or incomplete platforms. While our approach is centered on a commonly used genome-wide microarray (the Illumina Global Diversity Array), these same solutions can be translated to other settings.

## Research Design and Methods

### Regulatory Approval

The biobank at the Colorado Center for Personalized Medicine study data and protocol, including primary consents, was approved under Colorado Multiple Institutional Review Board #15-0461 and 20-2289, with additional approval for research operations via the CCPM Executive Committee. Analyses in UK Biobank were performed under primary approval 100578 with GDPR approval provided under the Colorado Multiple Institutional Review Board.

### Genetic Data in CCPM

To maximize our sample size with available overlapped genetic, demographic, and phenotypic information, we conducted this study using available data in CCPM^14,15^ (N=73,347) subset to variants found on the Illumina Global Diversity Array (GDA). We chose to perform our analyses on a joint set of individuals genotyped on GDA directly and individuals sequenced on an exome/genomic backbone platform to maximize statistical power. Genetic similarity is inferred with random forests using similarity to one of the six following superpopulation groups whose labels are defined in a publicly available reference panel (1000 Genomes Project and Human Genome Diversity Project): African (AFR), American (AMR), Central South Asian (SAS), East Asian (EAS), European (EUR), and Middle Eastern (MSE). Each individual in CCPM is assigned to a single genetic similarity group using group labels defined in the reference panel (Supplemental Figure 1). CCPM genetic similarity groups are labeled according to the following formula: “reference group”-like (e.g., African-like or European-like). For a detailed description of the data generation and quality control, see Rafaels et al. 2025, and for details of phecode phenotyping and covariate derivation, see Wiley et al. 2024.

We performed variant quality control using a pilot dataset of CCPM individuals genotyped using the GDA (N=48,634) to identify a subset of high-quality GDA variants to use for score calculation. To create this subset, we implemented a series of filters in a dataset consisting of 530 GDA plates. First, variants that did not harmonize with a well-established publicly available reference panel (TGP+HGDP)^16^ (either not present in the reference panel, indel variants, or mismatched alleles) were removed (n=610,879) using GenotypeHarmonizer^17^. Variants missing more than 1% of sample genotypes were removed (n=41,543). To identify variants that are not in Hardy-Weinberg equilibrium, six genetically homogeneous groups within CCPM were defined using the public reference panel (TGP+HGDP). Individuals were assigned to a group if their principal component coordinates fell within the space that encompasses 95% of a reference similarity group’s individuals across the first four principal components. Variants with a Hardy-Weinberg equilibrium p-value < 1×10^−8^ in any of the groups were excluded (n=17,311). Any variant with a plate batch effect p-value < 1×10^−12^ (Fisher’s exact test) was removed (n=78,148). Variants with a mean 10th percentile (by plate) GenCall score < 0.15 were removed (n=11,654). Variants with an 10th/90th-percentile LogR Ratio (LRR) score (used to detect copy number variation) substantially different from mean 10th/90th-percentile LRR score (mean +/-3 standard deviations in the case of 10th-percentile and mean +/-5 standard deviations in the case of 90th-percential scores) were removed (n=1,084). Variants with allele frequencies that differ substantially from population-matched Gnomad allele frequencies based on Mahalonobis distance were removed (n=45,936)^18^. Finally, any variant at a genomic position where the sum of the frequency of the two most frequent alleles is < 0.9999 (calculated using Gnomad v4) was removed (n=45,471) to avoid issues with tri-allelic misclassification. In total, 1,140,494 variants remained after filtering.

These SNP selection steps need only be performed one time but provide a suitable pool from which SNPs can be selected for future scores calculated with the GDA. In the absence of laboratory genotype validation of all variants, we used 4 control samples with known genotypes on each plate to test the efficacy of our SNP selection strategy. Genotype data at selected variants are significantly more concordant than those that were removed (mean concordance: 99.99% vs 99.09%, p<1e-16).

### Curated Diabetes Subtypes

Diabetes status was determined for individuals in CCPM using a clinician-curated algorithm based on integrating previously published classification methods^19–22^. General diabetes status was determined using a combination of diabetes diagnosis codes, broad diabetes medication use, and A1C from electronic medical records (Supplemental Figure 2). General diabetes cases were then further split into type 1 (Supplemental Figure 3) and type 2 cases (Supplemental Figure 4) based on the specific context of their diabetes diagnosis codes, available antibody test values, more granular medication use, and available lab values. The following analyses using CCPM participants focus on the curated subtype output from this algorithm (Supplemental Table 1).

### Score Calculation

We evaluated two established type 1 diabetes scores: 1) the 67-SNP GRS2^7^ developed in a European population which includes tagging variants to call HLA DR-DQ haplotypes, HLA variants, and non-HLA variants; and 2) a seven-SNP score developed in an African-descent population^12^ (AA7). GRS2 has previously been evaluated in multiple contexts (e.g.^23,24^) but the AA7 score is smaller and developed in an admixed-African setting, which may provide better portability across ancestry groups^25^.

Not all original variants in the published PRSs were present in the GDA subset that passed quality control checks (above). To capture these untyped variants, we identified proxy variants based on linkage disequilibrium (LD) (Supplemental Figure 5), leveraging the intrinsic local correlation structure present in the genome. For each biallelic variant, we searched for LD proxies in the 1000 Genomes^26^ reference populations using the LDproxy function in the LDlinkR package^27,28^. Seven proxy variants were lifted to grch38 using the liftOver R package^29,30^. From the list of potential proxy variants output by the LDproxy function, we removed SNPs that were not present in the quality controlled GDA subset or in the CCPM dataset. The remaining SNP with the highest R^2^ (i.e. in LD with the original variant) was selected as the best proxy SNP. If either the original variant or the selected proxy variant had ambiguous alleles (A/T or C/G), or LDproxy was unable to provide correlated alleles for the proxy variant, we verified the effect allele by matching the allele frequency of the original effect allele to the corresponding (major or minor) allele in the CCPM sample. For SNPs in the GRS2 used to call the HLA DR-DQ haplotypes, we additionally ensured that the original minor alleles aligned to the reference panel used in the GRS2 haplotype calculation pipeline correspond to the minor alleles (or its equivalent proxy alleles) in our samples (Supplemental Table 2). We identified proxy variants for 7 out of 7 SNPs of the AA7 score (mean R^2^ = 0.76) and 51 out of 67 SNPs of the GRS2 (mean R^2^ = 0.67) (Supplemental Table 3), with the rest captured directly on the GDA. For two SNPs of the AA7 score with poor proxy variants (R^2^ < 0.4), we tested removing those variants as an additional sensitivity analysis.

The full AA7 and the biallelic portion of the GRS2 were calculated using the ESCALATOR pipeline (https://github.com/menglin44/ESCALATOR), and the PRS Toolkit for HLA (https://github.com/USF-HII/hla-prs-toolkit) was used for calling HLA haplotypes using GRS2 tagging variants.

### Linear calibration of ancestry on PRS

To ensure that clinical applications of PRS can be beneficial for individuals across the continuous spectrum of ancestry, we calibrated scores for ancestry effects by linear adjustment of both the mean and variance of score distributions using the top 10 genetic PCs (Supplemental Figure 6-8), following methods described by the eMERGE Network^31,32^. This adjustment allows ancestry to vary on a continuous scale, rather than assigning individuals to discrete ancestry groups, better reflecting the reality of genetic ancestry variation.

By modeling PRS distribution being normally distributed, we can adjust ancestry’s effect from both the mean and variance –

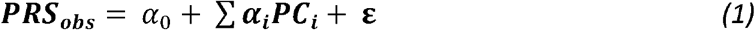

Where PRS_obs_ is the estimated PRS values that can be modeled with a linear relationship with PCs. The variance can be modeled similarly, leveraging the local correlations present in the genome

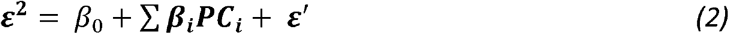

And ancestry-adjusted PRS can be calculated as

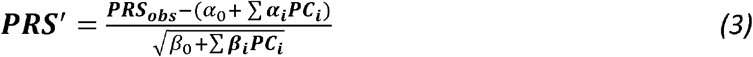

### Performance Evaluation

We evaluated performance of both proxy-variant scores as well as an additive combination of the two for discriminating between clinician-curated diabetes subtypes (type 1 vs type 2). Importantly, this “case-case” application is closer to the clinical setting of disease subtyping than the commonly reported comparison to healthy controls. For each score, the area under the receiver operating characteristic curve (AUC) was calculated using the pROC R package^33^. Model covariates included age, sex, genotyping batch, and the first 10 genetic PCs. We calculated AUC within genetically inferred ancestry groups to measure cross-ancestry performance. We tested pairwise comparisons of AUC across the three scores (GRS2, AA7, and their combination) using p-values calculated from the R2ROC^28^ (non-nested scores) and r2redux^29^ (nested scores) R packages.

We evaluated the strength of association between PRS and disease status, while accounting for covariates age, sex, genotyping batch, and the first 10 genetic PCs. We tested different thresholds for being considered “high risk” based on the top percentiles of PRS (top 5% - 25%) and calculated odds ratios (OR), sensitivity, and specificity metrics for each. Finally, to test across the adult age spectrum we performed sub-analyses stratified by age at first recorded diabetes diagnosis within groups <40 years, 40-60 years, and >60 years.

### Replication in UK Biobank

We replicated our score evaluation in the UK Biobank using ICD-10 code phenotypes for type 1 diabetes and type 2 diabetes, mapped from phecodes using the PheWAS R package^34^. Scores were evaluated in genetically defined ancestry groups from the PanUK Biobank^35^ that had more than 10 cases of type 1 diabetes. European-ancestry and African-ancestry populations passed this threshold for evaluation (Supplemental Table 1). Individuals with both type 1 diabetes and type 2 diabetes diagnosis codes were removed from the analysis.

### Additional Clinical Variables

Follow up analyses used additional clinical variables pulled from the biobank at CCPM including: 1) Glutamate decarboxylase (GAD) autoantibody testing, defined by LOINC code 56540-8 and considered to be antibody positive if any test value was above the reference range of 5.0 units/mL. 2) Diabetic ketoacidosis (DKA), defined by phecodes 250.11 or 250.21; 3) maximum A1C lab values, obtained by searching for strings “a1c” or “a1” in all available lab fields and then removing off-target matches (Supplemental Table 4-5); 4) insulin use, defined by searching for strings of insulin medication names in all available medication fields and removing off-target matches (Supplemental Table 6-7).

### Associations with Complications

To quantify the potential impact of clinical PRS testing, we evaluated the relationship between PRS and diabetic ketoacidosis (DKA). For those participants with DKA events, we stratified individuals by PRS decile and calculated the proportion of those with a DKA event in each bin. We similarly considered rates of GAD autoantibody testing and GAD autoantibody positivity by PRS decile. Finally, to help identify individuals with adult-onset disease, we further subset the data to investigate those with recorded antibody screening pre-DKA vs post-DKA.

## Results

### Participant characteristics

We used 69,515 unrelated individuals in CCPM, of which 1,186 had a type 1 diagnosis code in their EHR and 9,515 had a type 2 diabetes diagnosis code using an ICD-based phecode definition (937 individuals had codes for both subtypes). The curated phenotyping algorithm identified 892 individuals with type 1 diabetes and 7,255 individuals with type 2 diabetes. Individuals with diabetes in CCPM were 54.4% men; 77.1%, 5.3%, and 10.3% non-Hispanic white, non-Hispanic Black, and Hispanic race or ethnicity, respectively; and mean age was 59.7 years. Those with type 1 diabetes were younger (mean age = 49.8 years, p < 2.2×10^−16^) and had a higher proportion of non-Hispanic white (84.1%, p = 1.6×10^−9^) than those with type 2 diabetes (mean age = 63.7 years, 74.9% non-Hispanic white). Of those with a type 2 diabetes diagnosis code, 637 were reclassified as type 1 diabetes when evaluated by our clinician curated algorithm (Figure 1).

**Figure 1.**
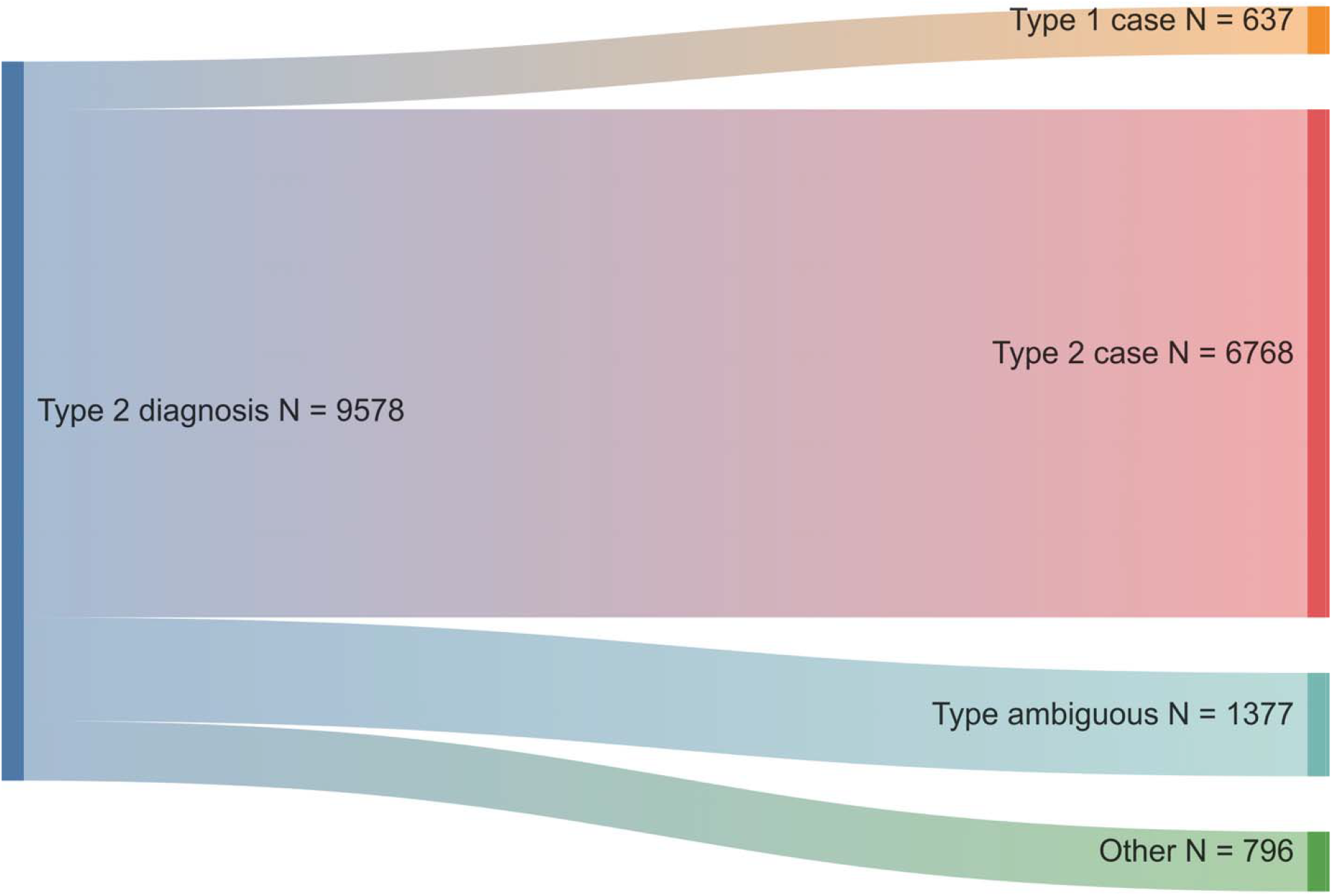
Individuals with type 2 diabetes diagnosis codes were classified by our clinician curated algorithm as type 1 diabetes cases, type 2 diabetes cases, type ambiguous diabetes, or other (non-diabetic or prediabetic).

### PRS Performance Evaluation

All three evaluated proxy scores (GRS2, AA7, and an additive combination of the two) strongly differentiated between individuals with type 1 diabetes and individuals with type 2 diabetes when adjusting for demographic covariates (Figure 2A, Supplemental Figure 9-10). In CCPM, scores were evaluated in groups of individuals with high genetic similarity to continental ancestry groups in the reference panel (see Methods) and that had >10 type 1 diabetes cases, including EUR-like, AFR-like, AMR-like, and EAS-like genetic similarity groups (Supplemental Table 8). Across all genetic similarity groups, the scores performed similarly well (AUC > 0.80) and most AUC estimates had substantially overlapping confidence intervals. The largest difference in AUC between the AA7 and the GRS2 was among EUR-like individuals; the GRS2 had the strongest predictive performance (AUC = 0.87 [95% CI 0.86-0.89]), but the simpler AA7 score performed nearly as well (AUC = 0.82 [95% CI 0.81 – 0.84]). We tested for significant differences in score performance between scores within each genetic similarity group (Supplemental Table 9), and although some AUC values were significantly different (p < 0.05), particularly in the EUR-like group, all scores performed well across all groups.

**Figure 2.**
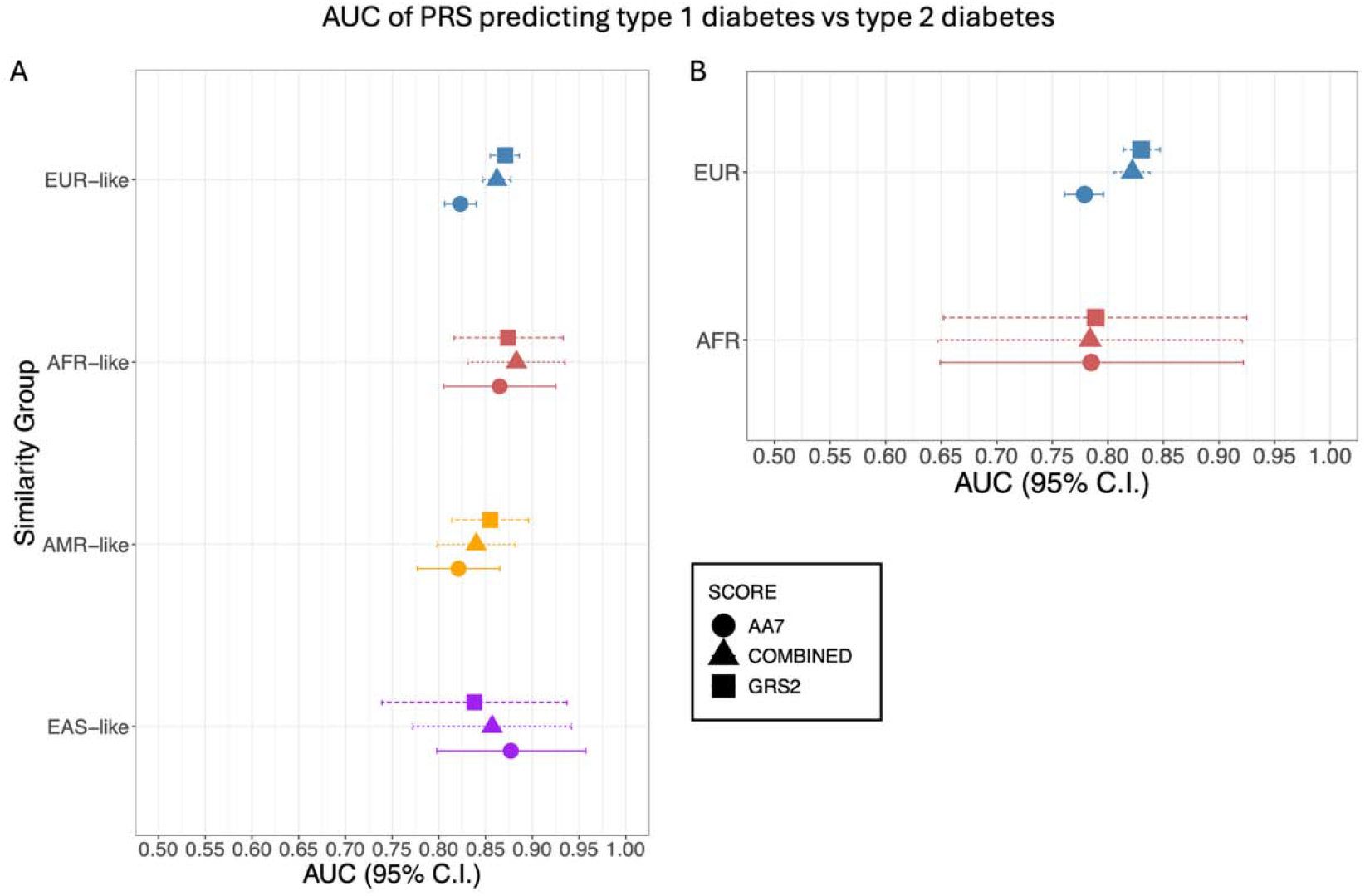
Performance of PRS models in differentiating clinician curated diabetes subtypes in CCPM (A) or ICD-10 code diabetes subtypes in the UK Biobank (B). Models included covariates: age, sex, genotyping batch, and the top 10 genetic PCs within each genetically defined similarity group containing more than 10 type 1 diabetes cases. Model performance is quantified using the area under the receiver-operator curve (AUC) as a measurement of classifier accuracy.

Predictive performance of all scores was similarly strong when evaluated in the UK Biobank in EUR-like and AFR-like groups (Figure 2B), despite using diabetes classification from diagnosis codes only. In the EUR-like group, the GRS2 had the best predictive performance (AUC = 0.83 [95% CI 0.81-0.85]), although the AA7 also performed well (AUC = 0.78 [95% CI 0.76-0.80]. In the AFR-like group, the three scores had nearly identical estimates (AUC = 0.78-0.79, Supplemental Tables 10-11).

Score performance based on AUC was not sensitive to removing the lowest-LD proxy variants from the AA7 score (Supplemental Tables 12-13). Similarly, AA7 performance for discriminating type 1 and type 2 diabetes was not impacted by including a T2D PRS^36^ in the model (p = 0.12-0.82) for each ancestry group, Supplemental Tables 14-15). Finally, we evaluated performance of PRS stratified by age at first diabetes diagnosis and found that scores better predicted type 1 diabetes in groups with lower age-at-diagnosis across ancestry groups (Supplemental Figure 11, Supplemental Tables 16-18). We stratified by age-at-diagnosis using groups <40, 40-60, and >60. For EUR-like individuals, AUC values ranged from 0.87 [95% CI 0.84-0.89] in the youngest age group down to 0.71 [95% CI 0.65-0.76] in the oldest age group. Similarly, for AFR-like individuals, AUC was highest in the youngest group (0.91 [95% CI 0.85-0.97] in those under 40). This is consistent with our expectations as younger individuals have had less environmental exposure contributing to disease onset, thereby revealing a greater genetic contribution.

Given the simplicity of the AA7 score compared to the GRS2 (both in number of SNPs and the process of calling of HLA DR-DQ haplotypes), and its strong performance across ancestry groups in differentiating type one diabetes cases from type 2 diabetes cases, it represents a realistic and clinically useful option for clinical PRS application. Our subsequent analyses highlight results from the AA7 score due to this simplicity, although we note that our released pipeline can leverage either scoring regime.

To evaluate whether there is potential for a useful clinical risk threshold for the AA7 score, we compared the risk of type 1 diabetes diagnosis for those with the highest AA7 scores versus the rest of the population, while accounting for covariates. When classifying high-risk of type 1 diabetes within each ancestry similarity group using the top 25%, 20%, 15%, and 5% of the PRS distribution, the odds ratios of type 1 diabetes in our largest group (EUR-like) were 3.66 [95% CI 3.08-4.34], 4.46 [95% CI 3.74-5.31], 5.13 [95% CI 4.28-6.15], and 3.96 [95% CI 3.05-5.15](Table 1, Supplemental Table 19).

**Table 1.** Evaluation of thresholds for determining individuals genetically “at-risk” for type 1 diabetes based on the ancestry-calibrated AA7 PRS. Individuals in the top percentiles of the score have increased odds of having type 1 diabetes compared to those below the score percentile cutoff. OR = odds ratio, CI = 95% confidence interval, PPV = positive predictive value, NPV = negative predictive value.

| *Ancestry* | *Percentile* | *OR* | *CI Lower* | *CI Upper* | *P* | *Sensitivity* | *Specificity* | *PPV* | *NPV* |
| --- | --- | --- | --- | --- | --- | --- | --- | --- | --- |
| *AFR-like* | 95% | 6.42 | 2.43 | 16.97 | 1.78E-04 | 0.74 | 0.53 | 0.96 | 0.12 |
|  | 90% | 4.48 | 1.84 | 10.91 | 9.57E-04 | 0.74 | 0.53 | 0.96 | 0.12 |
|  | 85% | 4.29 | 1.88 | 9.79 | 5.36E-04 | 0.74 | 0.53 | 0.96 | 0.12 |
|  | 80% | 3.66 | 1.64 | 8.15 | 1.52E-03 | 0.74 | 0.53 | 0.96 | 0.12 |
|  | 75% | 2.26 | 1.03 | 4.96 | 4.15E-02 | 0.74 | 0.53 | 0.96 | 0.12 |
| *AMR-like* | 95% | 2.40 | 1.09 | 5.29 | 3.04E-02 | 0.73 | 0.46 | 0.94 | 0.14 |
|  | 90% | 2.40 | 1.34 | 4.29 | 3.17E-03 | 0.73 | 0.46 | 0.94 | 0.14 |
|  | 85% | 2.28 | 1.35 | 3.85 | 1.97E-03 | 0.73 | 0.46 | 0.94 | 0.14 |
|  | 80% | 2.47 | 1.50 | 4.07 | 4.08E-04 | 0.73 | 0.46 | 0.94 | 0.14 |
|  | 75% | 2.79 | 1.72 | 4.52 | 3.21E-05 | 0.73 | 0.46 | 0.94 | 0.14 |
| *EAS-like* | 95% | 55.55 | 3.36 | 919.27 | 5.02E-03 | 0.66 | 0.47 | 0.93 | 0.12 |
|  | 90% | 7.17 | 1.06 | 48.57 | 4.35E-02 | 0.66 | 0.47 | 0.93 | 0.12 |
|  | 85% | 2.57 | 0.56 | 11.77 | 2.25E-01 | 0.66 | 0.47 | 0.93 | 0.12 |
|  | 80% | 2.47 | 0.61 | 9.96 | 2.03E-01 | 0.66 | 0.47 | 0.93 | 0.12 |
|  | 75% | 2.99 | 0.70 | 12.85 | 1.40E-01 | 0.66 | 0.47 | 0.93 | 0.12 |
| *EUR-like* | 95% | 3.96 | 3.05 | 5.15 | 7.83E-25 | 0.74 | 0.57 | 0.93 | 0.24 |
|  | 90% | 5.65 | 4.65 | 6.88 | 4.92E-67 | 0.74 | 0.57 | 0.93 | 0.24 |
|  | 85% | 5.13 | 4.28 | 6.15 | 3.11E-70 | 0.74 | 0.57 | 0.93 | 0.24 |
|  | 80% | 4.46 | 3.74 | 5.31 | 3.28E-63 | 0.74 | 0.57 | 0.93 | 0.24 |
|  | 75% | 3.66 | 3.08 | 4.34 | 1.34E-49 | 0.74 | 0.57 | 0.93 | 0.24 |

### Score integration with diabetes-related clinical variables

We performed association testing between score predictions and additional clinical evidence, including records of confirmatory tests of GAD autoantibodies and DKA with scores in deciles. We found that the top quintile of the ancestry-calibrated AA7 score showed significant enrichment for GAD antibody test positivity and DKA (Figure 3A-B). Of available GAD autoantibody tests, 42.8% of test positivity occurred in those in the top 20% of the AA7 score, even though fewer than 2% of individuals received GAD antibody testing. Additionally, nearly half (48%) of DKA cases in our sample occurred in individuals in the top 20% of genetic risk for type 1 diabetes based on the AA7 score. Among individuals with a DKA event and GAD antibody testing occurring concurrently with or after the DKA event, 38% were in the top quintile of the AA7 score, suggesting that genetically informed early antibody testing could help avoid this severe diabetes complication.

**Figure 3.**
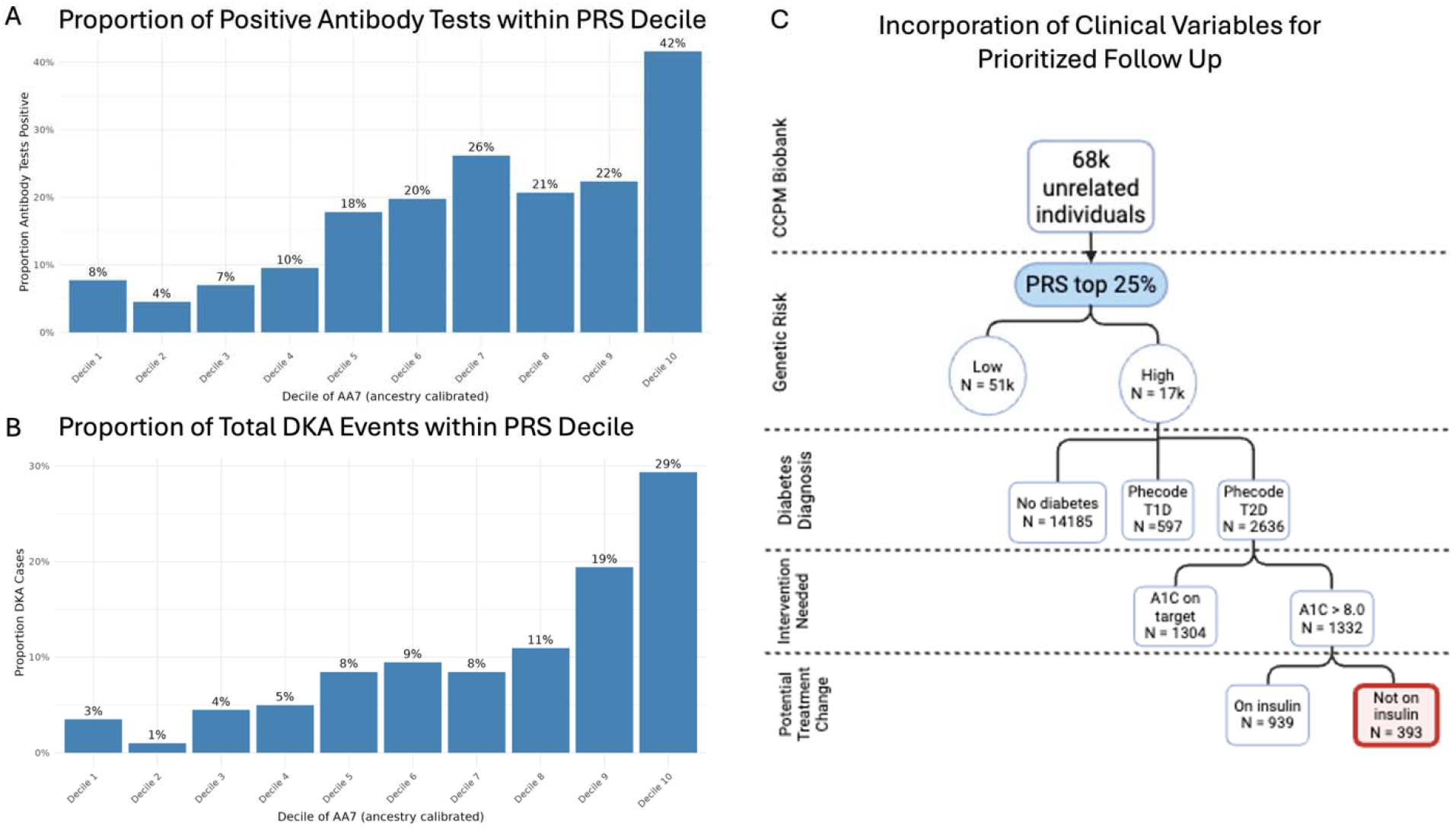
Relationships between ancestry-calibrated AA7 PRS decile and diabetes-relevant clinical variables. A) The proportion of individuals with a GAD autoantibody test that had a positive result for each decile of the ancestry-calibrated AA7 score (Number of positive tests in decile / Number of total tests in decile). There were 929 individuals in CCPM with a GAD antibody test. B) The proportion of total DKA cases that occurred in each decile of the ancestry-calibrated AA7 score (Number of DKA cases in decile / Number of DKA cases total). There were 201 individuals in CCPM with a DKA event. C) Multiple criteria are combined with PRS to identify population of interest for follow-up. We identified a population of 393 individuals who could benefit from a clinical PRS test for type 1 diabetes risk. These are individuals with evidence of genetic risk for type 1 diabetes (in the top 25% of the PRS), with a type 2 diabetes diagnosis code, evidence of need for additional intervention for disease management, and not already on insulin.

Based on criteria for an at-risk population, in consultation with our clinical experts, we combined genetic risk, current diabetes status, and relevant clinical variables from health records to identify a proposed population of interest in CCPM that could benefit from additional testing for type 1 diabetes (Figure 3C) out of our entire population. We used the top 25% of the AA7 score as a threshold here for this proof-of-concept study due to similar effect sizes across ancestries but acknowledge that a final risk threshold for a clinical PRS test should be determined in collaboration with clinical and health system experts. Nonetheless, using this specific threshold, we found that there were 393 individuals in the CCPM biobank whose clinical records included only type 2 diagnosis codes, had at least one A1c >8.0, and had no history of insulin treatment. Additional future investigations are warranted to follow up with this group as well as determine an optimal threshold balancing sensitivity, specificity, and clinical feasibility.

## Conclusions

We have developed a robust framework integrating analytically valid measures of type 1 diabetes polygenic scoring into a clinical/translational setting, with a specific use case for aiding in the classification of adult-onset autoimmune diabetes (LADA). We adapted well-established type 1 diabetes scores and SNPs directly genotyped on a major array platform for this approach. We have made code for proxy SNP selection and ancestry calibration available as a substantial step forward in the path toward the clinical use of PRS.

Our analyses focus on type 1 diabetes as a proof-of-concept for PRS clinical utility, and we demonstrate that the previously established, small seven-SNP AA7 score performs well in differentiating type 1 diabetes cases from type 2 diabetes cases in both the CCPM and UK biobanks. The score performs nearly as well as the more complex GRS2, which has 67 SNPs and requires using tagging variants to call specific HLA DR-DQ haplotypes. This is particularly important for the practicality of any clinical implementation, since a smaller score significantly eases the burden of analytic validation: for multianalyte algorithmic analyses (like PRS) with a limited number of contributing genetic variants, full analytic validation of the accuracy of the genotyping or sequencing assay at each locus should be performed, because the weight of each variant to the PRS outcome is relatively large. Non-clinical applications supplementing or contextualizing PRS (e.g., ancestry estimation) may be performed without the same level of analytic or clinical validation. This significantly empowers clinical labs to work with PRS with small numbers of variants while incorporating context from the rest of the genome as with ancestry calibration. As we describe here, our ancestry estimation still was shown to be robust, even under higher levels of missingness allowed by our QC process.

Importantly, the AA7 score performs well across all genetic ancestry groups that were represented in our analyses, which is essential for genetic risk to be evaluated equitably in a clinical setting. Clinical use of PRS necessarily to identify a “high-risk” group requires determining a relevant threshold. Using a single score threshold for all individuals can disproportionally assign risk to specific groups confounded by genetic ancestry^37^. We demonstrate a potential solution to this problem for future scores, following a similar approach to the eMERGE consortium^31,32^. This is in-line with guidance from the March 2023 NASEM report^38^ recommendations for continuous ancestry estimation, rather than classifying individuals into discrete groups, which may not truly represent the full spectrum of human genetic diversity.

Another notable advantage of integrating polygenic risk scores (PRS) into clinical diagnostics is the potential to reduce avoidable healthcare costs. For instance, the diagnosis of diabetic ketoacidosis (DKA) typically necessitates hospital admission for continuous monitoring and treatment, which incurs an average cost of approximately $30,836 per admission^39,40^, with an average length of stay of 3.02 days^41^. Admissions for DKA cost the US healthcare system $6.76 billion in 2017^39^. If a substantial proportion of these cases could be prevented through early identification of type 1 diabetes—aided by PRS and confirmatory antibody testing—timely initiation of effective treatments could substantially mitigate the high costs associated with downstream health care utilization.

Despite our success developing a framework for clinical implementation of type 1 diabetes PRS, challenges lie ahead for other diseases and complex genetic risk scores. First, as the field advances in developing more robust polygenic risk scores, many newer and better scores incorporate millions of markers, creating additional challenges in clinical validation. This will likely call for developing additional steps to create sparser scores. Second, although ancestry-recalibration of PRS is efficient thanks to the use of static measures of continuous ancestry, the effects of missing/inaccurate genotypes during PRS recalibration must be evaluated for each score independently within the context of the clinically relevant PRS threshold under consideration.

Finally, we note the AUCs herein are slightly lower than in other studies, particularly those with a type 1 diabetes focus^7^. This reflects multiple sources of noise in our large adult-focused biobank, including: the necessity of directly-typed proxy variants, disease heterogeneity in a large-scale US health system, and the limitations of measuring a disease with lower heritability (and thus expected lower PRS performance) in later-life settings. This same trend of higher heritability in early-onset conditions has been found in prior research into adult-onset type 1 diabetes^42^ as well as other complex traits^43^, and reflects the lower cumulative environmental burden earlier in life. Consistent with this assumption, stratifying our analysis on age showed the strongest AUC in lower-age individuals (<40, when compared to 40-60 and 60+).

Overall, our results demonstrate the utility of PRS for aiding in diabetes subtyping, and our pipeline opens the door for evaluation of additional PRS for clinical application in the future. We recommend prioritizing scores for potential clinical implementation in diabetes and related traits based on the following criteria: (1) a strong genetic basis (i.e. high narrow-sense heritability), (2) presence of well-documented, peer-reviewed PRS models with significant associations to the trait(s) of interest, and (3) potential for achieving effective and equitable intervention without unintentionally exacerbating disparities. Adult-onset type 1 diabetes remains a highly promising candidate for PRS implementation due to all these factors.

Early and consistent collaboration with clinical domain experts should be a prerequisite for any further PRS implementation efforts to ensure that PRS can be applied in a clinically relevant and responsible manner. Our scoring approach for adult-onset diabetes has proven to be highly impactful and should not be limited to our site. To that end we provide a code repository for following this same approach for other platforms, ensuring our approach to variant quality control, proxy variant detection, and ancestry recalibration can be available to all. Our final scores, generated in a clinical lab, demonstrate high clinical efficacy and impact. This combined with antibody screening brings us closer to a future with effective use of clinical polygenic scoring for diabetes subtyping, ultimately reducing complications, disease misclassification, and healthcare utilization.

## Data Availability

Summary statistics for the project are contained in the manuscript. Additional code and resources available upon reasonable request to the authors. Due to patient privacy/consent, individual-level data of the CCPM biobank is not available.

## Acknowledgements

We are deeply indebted to the participants in CCPM, without whom this work would not be possible. We acknowledge Regeneron Genetics Center and the Colorado Center for Personalized Medicine for making the data available for this project. We thank members of the CCPM community for helpful feedback on presentations on this project. This work was partially supported by the Health Data Compass Data Warehouse project and the Colorado Center for Personalized Medicine. MSB, LKW, SR, ML, and CRG were additionally supported by R01HL151152. The opinions expressed in this article are the authors’ own and do not reflect the view of the National Institutes of Health, the Department of Health and Human Services, or the United States government.

## Author Contributions

CRG and ML designed the project with input from MSB, CSW, and JAS. Analyses were performed by MSB, MJF, JAS, NMR. Additional scientific and clinical feedback were provided by EK, KRC, RKJ, SO, TO, JBC, LKW, SR, and NR. KM and IMB facilitated access to EHR data in consultation with LKW and SR. The primary draft was written by MSB, SR, NR, ML, and CRG, with input and approval from all coauthors. ML and CRG supervised all portions of analysis. CRG is the guarantor of the work and had access to the full data in the study and takes responsibility for the accuracy of the analyses described.

## References

1. Khera, A. V. et al. Genome-wide polygenic scores for common diseases identify individuals with risk equivalent to monogenic mutations. Nat. Genet. 50, 1219–1224 (2018).

2. Menke, A. et al. The Prevalence of Type 1 Diabetes in the United States: Epidemiology 24, 773–774 (2013).

3. Bullard, K. M. et al. Prevalence of Diagnosed Diabetes in Adults by Diabetes Type — United States, 2016. MMWR Morb. Mortal. Wkly. Rep. 67, 359–361 (2018).

4. de Lusignan, S. et al. Miscoding, misclassification and misdiagnosis of diabetes in primary care. Diabet. Med. J. Br. Diabet. Assoc. 29, 181–189 (2012).

5. Pang, H. et al. The missing heritability in type 1 diabetes. Diabetes Obes. Metab. 24, 1901– 1911 (2022).

6. Oram, R. A. et al. A Type 1 Diabetes Genetic Risk Score Can Aid Discrimination Between Type 1 and Type 2 Diabetes in Young Adults. Diabetes Care 39, 337–344 (2015).

7. Sharp, S. A. et al. Development and Standardization of an Improved Type 1 Diabetes Genetic Risk Score for Use in Newborn Screening and Incident Diagnosis. Diabetes Care 42, 200–207 (2019).

8. Graham, J. et al. Genetic Effects on Age-Dependent Onset and Islet Cell Autoantibody Markers in Type 1 Diabetes. Diabetes 51, 1346–1355 (2002).

9. Pihoker, C., Gilliam, L. K., Hampe, C. S. & Lernmark, Å. Autoantibodies in Diabetes. Diabetes 54, S52–S61 (2005).

10. Feeney, S. J. et al. Evaluation of ICA512As in Combination With Other Islet Cell Autoantibodies at the Onset of IDDM. Diabetes Care 20, 1403–1407 (1997).

11. Yang, P. K. et al. Type 1 Diabetes Genetic Risk in 109,954 Veterans With Adult-Onset Diabetes: The Million Veteran Program (MVP). Diabetes Care 47, 1032–1041 (2024).

12. Onengut-Gumuscu, S. et al. Type 1 Diabetes Risk in African-Ancestry Participants and Utility of an Ancestry-Specific Genetic Risk Score. Diabetes Care 42, 406–415 (2019).

13. Ben-Eghan, C. et al. Don’t ignore genetic data from minority populations. Nature 585, 184– 186 (2020).

14. Wiley, L. K. et al. Building a vertically integrated genomic learning health system: The biobank at the Colorado Center for Personalized Medicine. Am. J. Hum. Genet. 111, 11–23 (2024).

15. Rafaels, N. et al. CCPM Freeze3 Materials and Methods. Zenodo 10.5281/ZENODO.15330578 (2025).

16. Koenig, Z. et al. A harmonized public resource of deeply sequenced diverse human genomes. Genome Res. 34, 796–809 (2024).

17. Deelen, P. et al. Genotype harmonizer: automatic strand alignment and format conversion for genotype data integration. BMC Res. Notes 7, 901 (2014).

18. White, S. L. et al. Global multi-ancestry genetic study elucidates genes and biological pathways associated with thyroid cancer and benign thyroid diseases. Preprint at 10.1101/2025.05.15.25327513 (2025).

19. Wilson, M. P. et al. 331 Reusing EHR Phenotyping Algorithms in Practice: Developing the Colorado Diabetes EHR Research Repository (CODER). J. Clin. Transl. Sci. 7, 98–99 (2023).

20. Kho, A. N. et al. Use of diverse electronic medical record systems to identify genetic risk for type 2 diabetes within a genome-wide association study. J. Am. Med. Inform. Assoc. 19, 212–218 (2012).

21. Schroeder, E. B., Donahoo, W. T., Goodrich, G. K. & Raebel, M. A. Validation of an algorithm for identifying type 1 diabetes in adults based on electronic health record data. Pharmacoepidemiol. Drug Saf. 27, 1053–1059 (2018).

22. Upadhyaya, S. G. et al. Automated Diabetes Case Identification Using Electronic Health Record Data at a Tertiary Care Facility. Mayo Clin. Proc. Innov. Qual. Outcomes 1, 100–110 (2017).

23. Cone Health. Helix and Cone Health Launch Population Genomics Initiative. https://www.conehealth.com/news/news-search/2023-news-releases/helix-and-cone-health-launch-population-genomics-initiative/ (2023).

24. Guertin, K. A. et al. Implementation of type 1 diabetes genetic risk screening in children in diverse communities: the Virginia PrIMeD project. Genome Med. 16, 31 (2024).

25. Lin, M., Park, D. S., Zaitlen, N. A., Henn, B. M. & Gignoux, C. R. Admixed Populations Improve Power for Variant Discovery and Portability in Genome-Wide Association Studies. Front. Genet. 12, 673167 (2021).

26. Auton, A. et al. A global reference for human genetic variation. Nature 526, 68–74 (2015).

27. Myers, T. A., Chanock, S. J. & Machiela, M. J. LDlinkR: An R Package for Rapidly Calculating Linkage Disequilibrium Statistics in Diverse Populations. Front. Genet. 11, 157 (2020).

28. Momin, Md. M., Wray, N. R. & Lee, S. H. R2ROC: An efficient method of comparing two or more correlated AUC from out-of-sample prediction using polygenic scores. Preprint at 10.1101/2023.08.01.551571 (2023).

29. Momin, M. M., Lee, S., Wray, N. R. & Lee, S. H. Significance tests for R2 of out-of-sample prediction using polygenic scores. Am. J. Hum. Genet. 110, 349–358 (2023).

30. Bioconductor Package Maintainer. liftOver: Changing genomic coordinate systems with rtracklayer::liftOver. (2024).

31. Khera, A. V. et al. Whole-Genome Sequencing to Characterize Monogenic and Polygenic Contributions in Patients Hospitalized With Early-Onset Myocardial Infarction. Circulation 139, 1593–1602 (2019).

32. Khan, A. et al. Genome-wide polygenic score to predict chronic kidney disease across ancestries. Nat. Med. 28, 1412–1420 (2022).

33. Robin, X. et al. pROC: an open-source package for R and S+ to analyze and compare ROC curves. BMC Bioinformatics 12, 77 (2011).

34. Carroll, R. J., Bastarache, L. & Denny, J. C. R PheWAS: data analysis and plotting tools for phenome-wide association studies in the R environment. Bioinformatics 30, 2375–2376 (2014).

35. Karczewski, K. J. et al. Pan-UK Biobank GWAS improves discovery, analysis of genetic architecture, and resolution into ancestry-enriched effects. 2024.03.13.24303864 Preprint at 10.1101/2024.03.13.24303864 (2024).

36. Pezzilli, S. et al. Contribution of rare variants in monogenic diabetes-genes to early-onset type 2 diabetes. Diabetes Metab. 48, 101353 (2022).

37. Martin, A. R. et al. Human Demographic History Impacts Genetic Risk Prediction across Diverse Populations. Am. J. Hum. Genet. 100, 635–649 (2017).

38. Committee on the Use of Race, Ethnicity, and Ancestry as Population Descriptors in Genomics Research et al. Using Population Descriptors in Genetics and Genomics Research: A New Framework for an Evolving Field. 26902 (National Academies Press, Washington, D.C., 2023). doi:10.17226/26902.

39. Ramphul, K. & Joynauth, J. An Update on the Incidence and Burden of Diabetic Ketoacidosis in the U.S. Diabetes Care 43, e196–e197 (2020).

40. Lyerla, R. et al. Recurrent DKA results in high societal costs – a retrospective study identifying social predictors of recurrence for potential future intervention. Clin. Diabetes Endocrinol. 7, 13 (2021).

41. Portillo-Canales, S., Peters, J., Francis-Morel, G. & Dhindsa, S. Disparities in Outcomes and Health Care Utilization for Diabetic Ketoacidosis Among Patients With Type 1 and Type 2 Diabetes Mellitus: A 6-Year National Retrospective Cohort Study. Endocr. Pract. 31, 784–789 (2025).

42. Wei, Y. et al. Familial aggregation and heritability of childhood-onset and adult-onset type 1 diabetes: a Swedish register-based cohort study. Lancet Diabetes Endocrinol. 12, 320–329 (2024).

43. Mostafavi, H. et al. Variable prediction accuracy of polygenic scores within an ancestry group. eLife 9, e48376 (2020).

