## Supplemental Figures for "Enabling reproducible type 1 diabetes polygenic risk scoring for clinical and translational applications"

FIGURE 1. PRINCIPAL COMPONENTS 1-3 IN CCPM BY GENETIC SIMILARITY GROUP 2
FIGURE 2. CLINICIAN ALGORITHM FOR GENERAL DIABETES DEFINITION 3
FIGURE 3. CLINICIAN ALGORITHM FOR TYPE 1 DIABETES DEFINITION 4
FIGURE 4. CLINICIAN ALGORITHM FOR TYPE 2 DIABETES DEFINITION 5
FIGURE 5. FLOWCHART OF PROXY SNP SELECTION FOR GRS2 6
FIGURE 6. EXAMPLE ANCESTRY CALIBRATION USING PRS FOR HEIGHT 7
FIGURE 7. CALIBRATION OF THE AA7 SCORE USING THE FIRST 10 GENETIC PRINCIPAL COMPONENTS 8
FIGURE 8. CALIBRATION OF THE GRS2 SCORE USING THE FIRST 10 GENETIC PRINCIPAL COMPONENTS 9
FIGURE 9. AA7 PRS DISTRIBUTION BY CLINICIAN-CURATED DIABETES STATUS 10
FIGURE 10. GRS2 PRS DISTRIBUTION BY CLINICIAN-CURATED DIABETES STATUS 11
FIGURE 11. SCORE PERFORMANCE STRATIFIED BY AGE GROUP 12

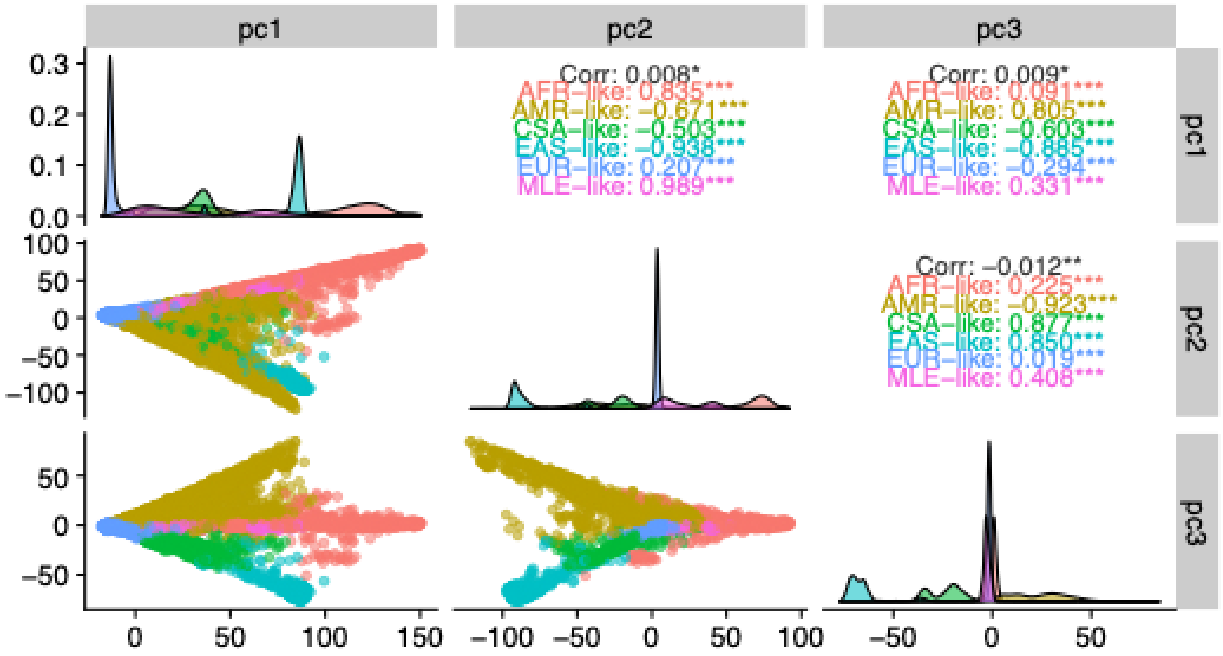


*Figure 1. Principal Components 1-3 in CCPM by genetic similarity group*

General DM
algorithm (Mayo)

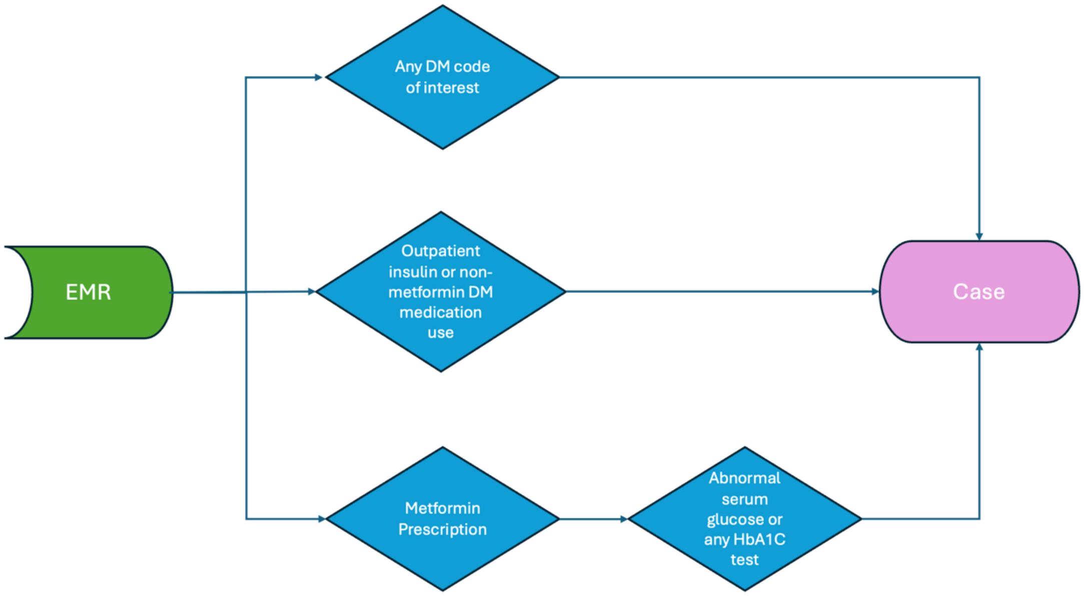


*Figure 2. Clinician algorithm for general diabetes definition*

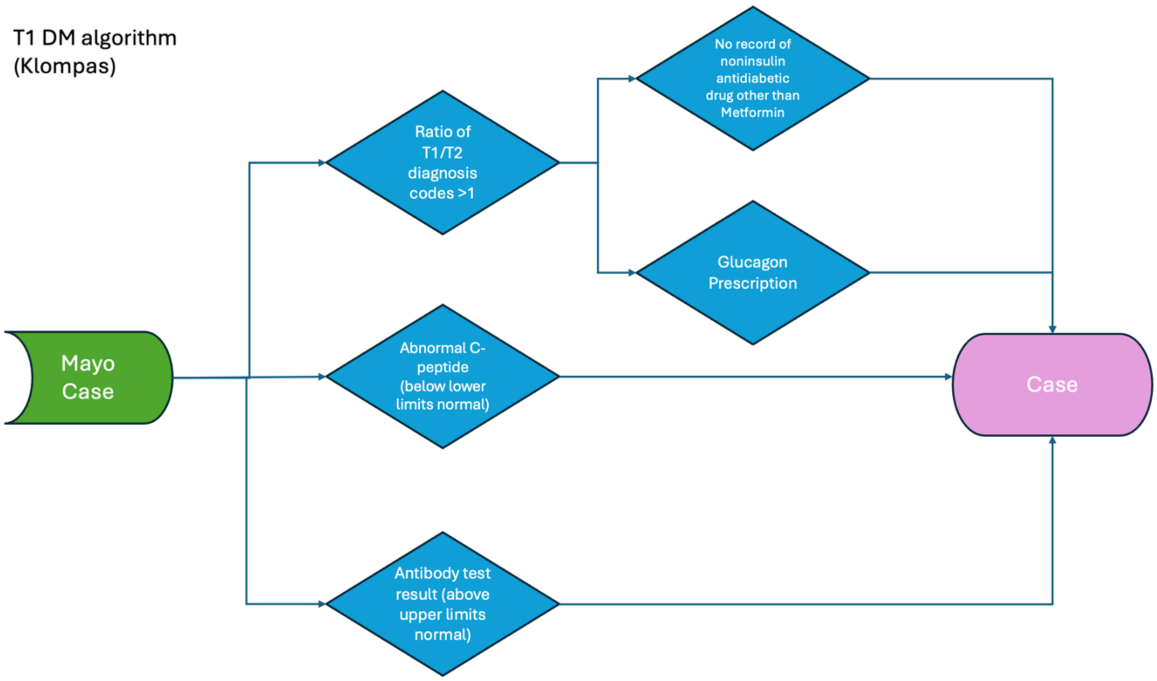


*Figure 3. Clinician algorithm for type 1 diabetes definition*

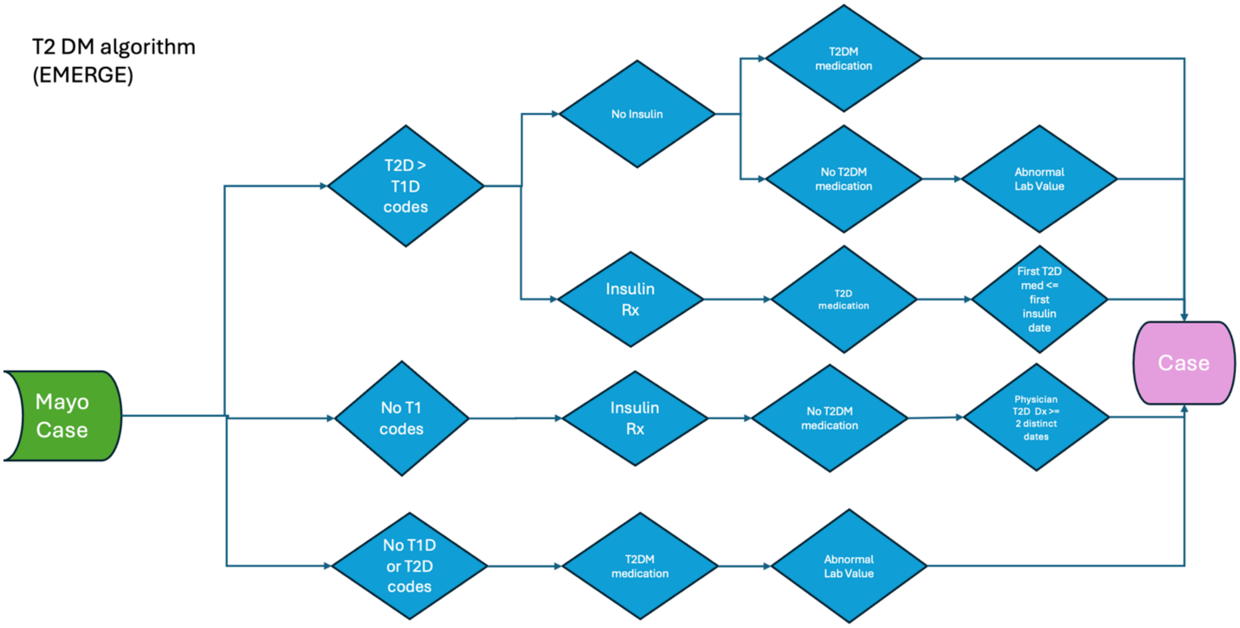


*Figure 4. Clinician algorithm for type 2 diabetes definition*

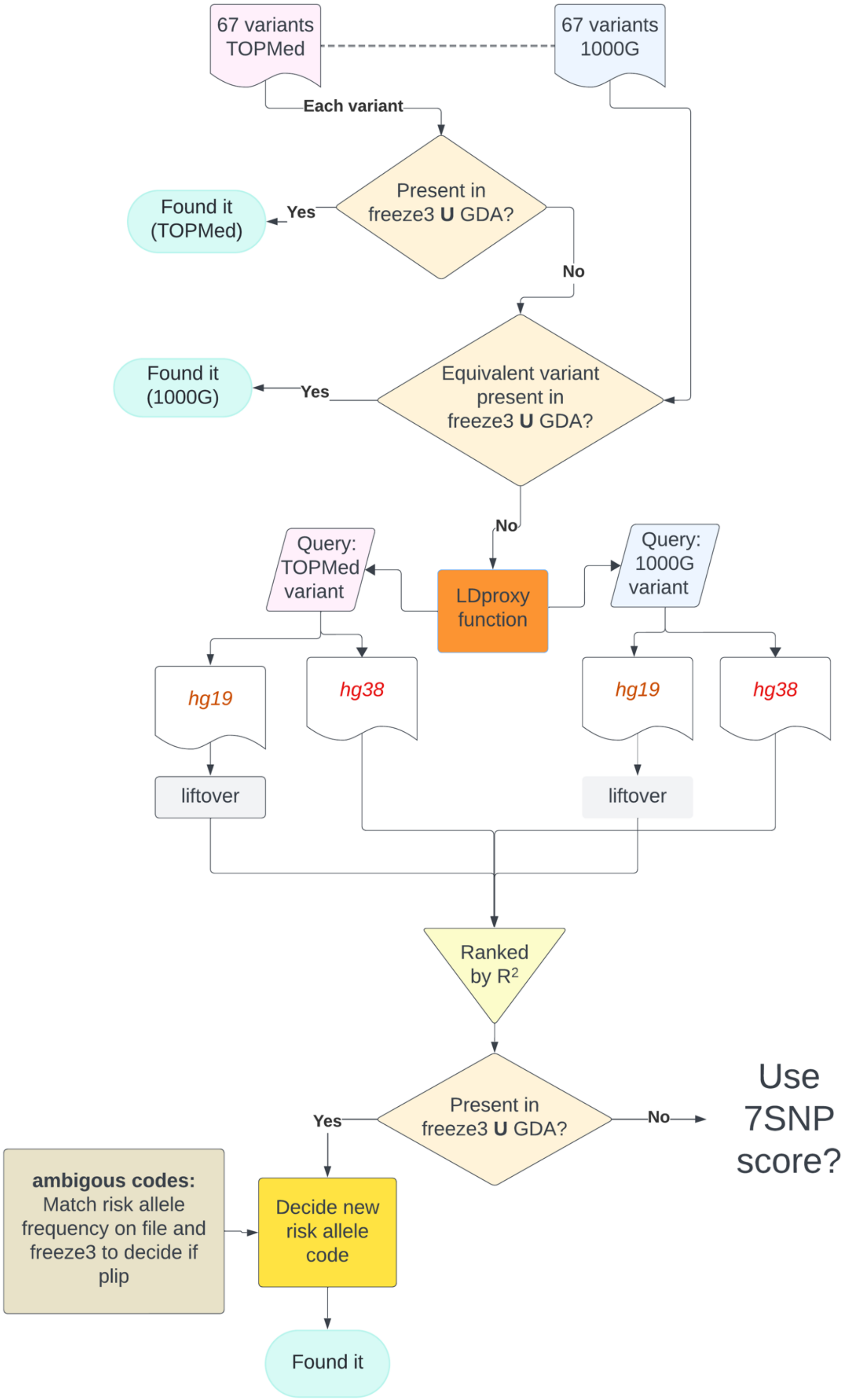


*Figure 5. Flowchart of proxy SNP selection for GRS2*

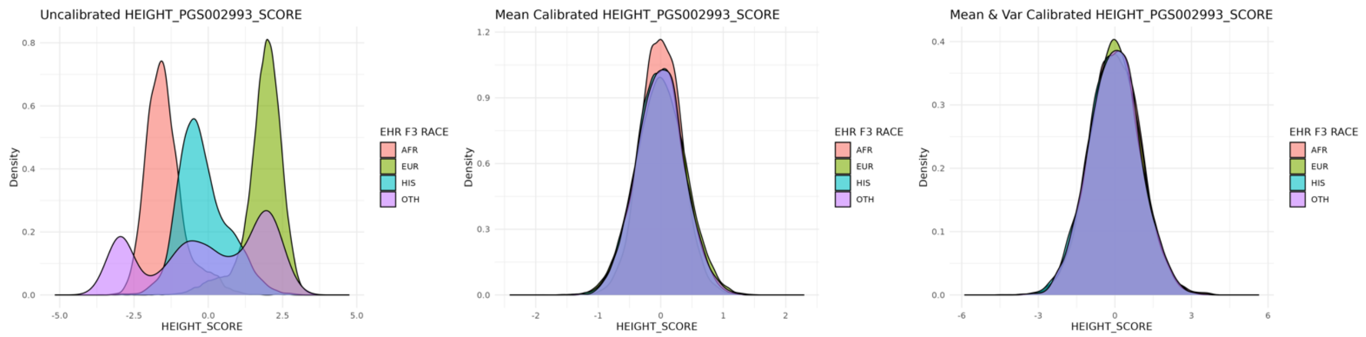


*Figure 6. Example ancestry calibration using PRS for height*

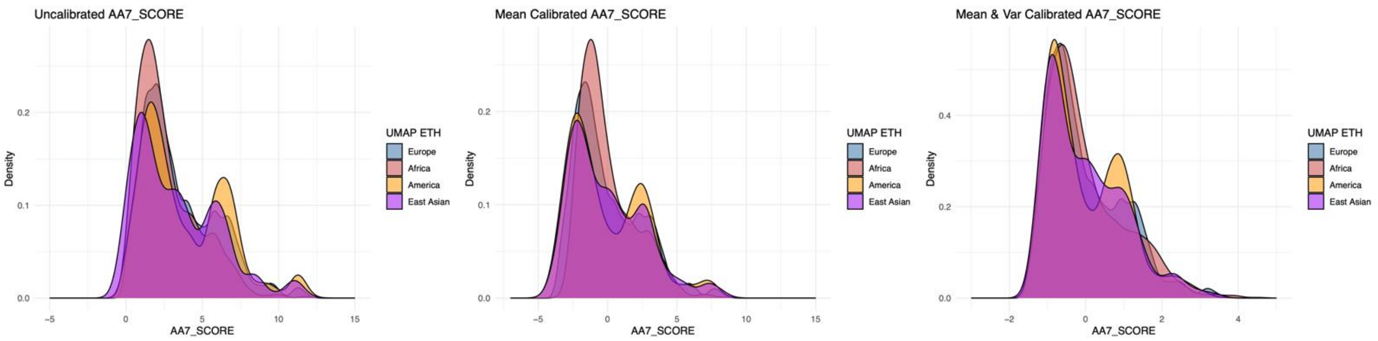


*Figure 7. Calibration of the AA7 score using the first 10 genetic principal components*

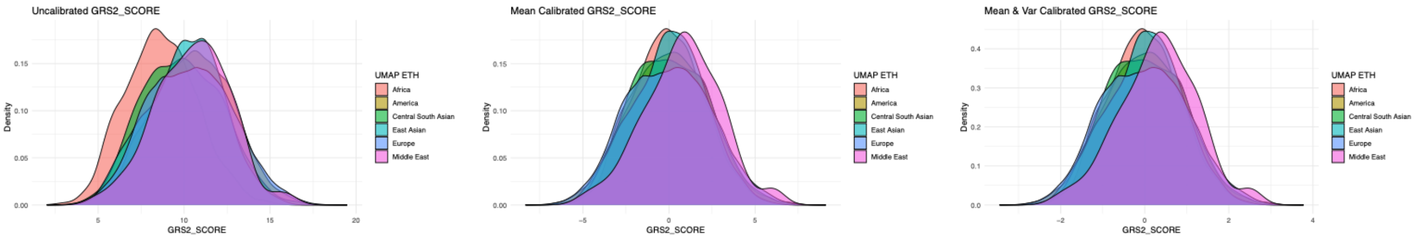


*Figure 8. Calibration of the GRS2 score using the first 10 genetic principal components*

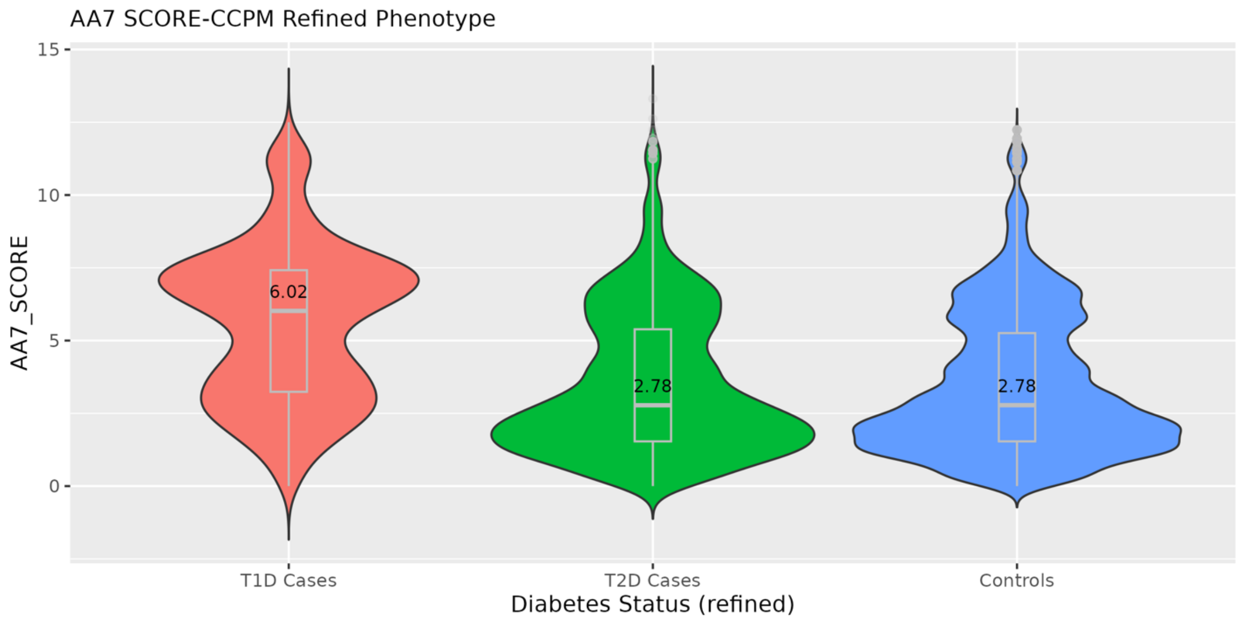


*Figure 9. AA7 PRS distribution by clinician-curated diabetes status*

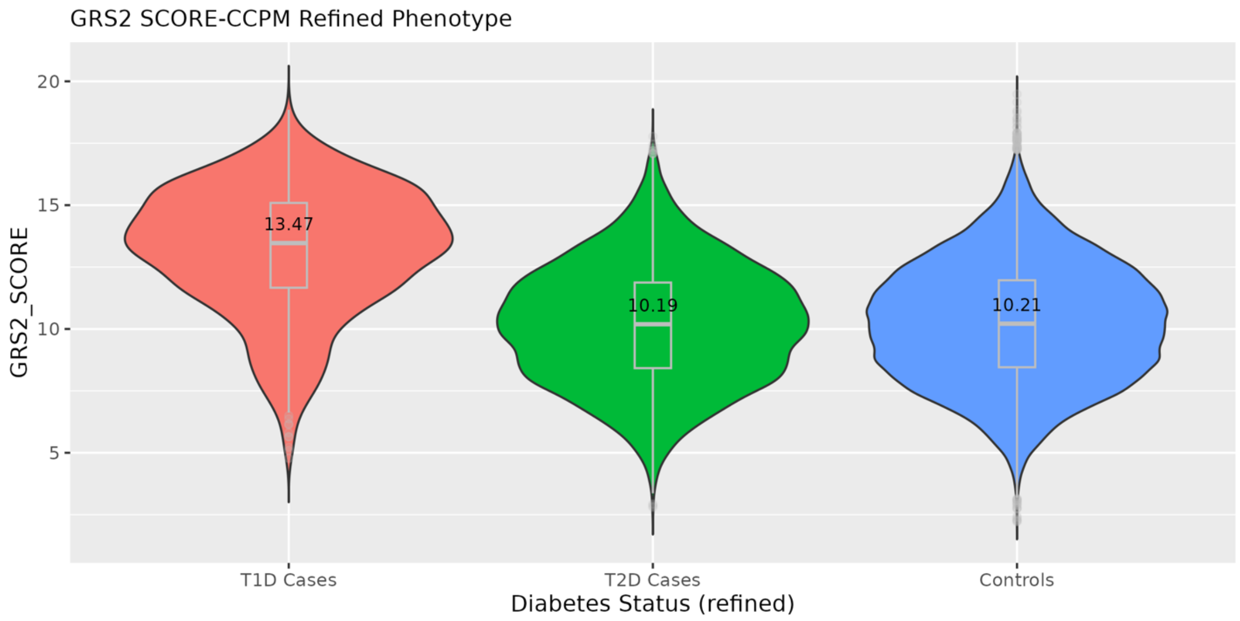


*Figure 10. GRS2 PRS distribution by clinician-curated diabetes status*

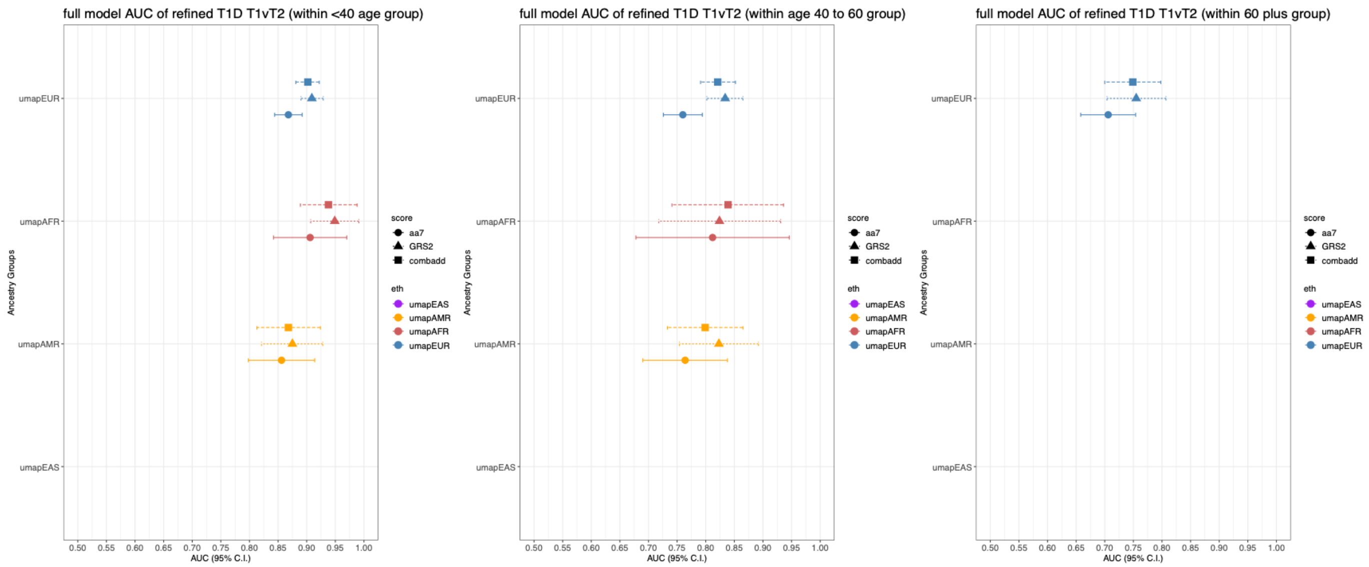


*Figure 11. Score performance stratified by age group*
